# Efficacy, Practicality, Intimal Health, and Sustainability of Menstrual Cups use among Undergraduate Medical Students: a cross-sectional study

**DOI:** 10.1101/2022.01.14.22269280

**Authors:** Brenda Czekalski Lobascz, Maria Beatriz de França Reis, Gabriella de Perez Monteiro e Tiburcio Mendes, Danillo Christian de Oliveira Cruz, Maria Jose Caetano Ferreira Damaceno, Carlos Izaias Sartorao Filho

## Abstract

**Introduction:** Women need effective, safe, and affordable menstrual hygiene products. The menstrual cup is an alternative method.

**Objective:** To identify the prevalence of menstrual cups among the medical students and the independent variables associated with their use adherence.

**Methods:** a cross-sectional study with an online survey applied to regularly matriculate women from FEMA Medical School in January 2021.

**Results:** Of 277 women, 164 participated. The mean age was 22.26 (SD 3.21). 136 preferred external pads, 60 internal pads, 28 menstrual cups, and 11 did not use. Using a 1-10 scale, price, efficacy, sustain, practice, intimal health status, hygiene, and importance of internal genitalia integrity were questioned. 37 (22,56%) women related the use of menstrual cups in the last year. Using any method to reduce menstruation had an odds ratio adjusted of 0.310 (95%CI 0.122-0.787). Concern about the environmental biodegradation had ORadj of 6.369 (95%CI 1.372-29.562); Intimal Health, ORadj of 1.996 (95%CI 1.183-3.368); internal genital integrity, ORadj of 0.824 (95%CI 0.682-0.995), for menstrual cups use.

**Conclusion:** women using a method to reduce their menstrual flow, concerning about biodegradation, concerning with intimal health, and with no concerns about manipulating their genitalia were significant independent factors for the adherence of menstrual cups.

## Introduction

Knowing women’s menstrual hygiene methods can tell us what they look for in a method and what they use as criteria to do so. The menstrual cup can be a viable option for most women during their menstrual period. (van Eijk et al. 2018) This method is not widespread in the population, and there is a lack of literature regarding the advantages and disadvantages of using it and the choosing criteria. (North and Oldham 2011; Smith et al. 2020) Sustainability is a current concern and can be one of the criteria adopted by women during their decision-making. (Jahan et al. 2020) Menstrual cups appear a sustainable, cost-effective, practical, and adequate choice compared to the most known methods. (Hennegan et al. 2020) Despite these observations, the use and the preference of menstrual cups remain primarily unexplored. (van Eijk et al. 2019)

We hypothesize that menstrual cups are underused among the population, even undergraduate medical women, who are supposed to be more knowledgeable about menstrual hygiene products.

Our primary objective was to identify independent factors in women predicting the preferences for using menstrual cups. In addition, the specific objective was to describe the prevalence of menstrual cups among undergraduate medical women.

## Methods

We used the STROBE guidelines to report this manuscript. (von Elm et al. 2008) The Faculty of Medicine of Educational Foundation of Municipality of Assis (FEMA), in São Paulo State, Brazil, provided human resources and infrastructure. We conducted a cross-sectional study approved by the FEMA Ethical Committee under the number 38034920.3.0000.8547, and all enrolled participants agreed to an informed written consent process before answering the online survey. Inclusion criteria: adult women regularly matriculated at FEMA Medical School. Exclusion criteria were women with previous pregnancies in the last 12 months. We calculate a sample size of 164 participants using the Slovin formula, based on the population size of 277 women regularly matriculated in the FEMA Medical course, a margin of error of 5%, a confidence interval of 95%, and an inaccurate standard deviation of 50%.

All participants were asked to answer a structured questionnaire to collect epidemiological and clinical data. This included: age in years, Caucasian or non-Caucasian, religion as Catholic, Protestants, other religions and non-religion, conjugal status as married or not married. The menstrual patterns included having an irregular menstrual cycle, a menstrual period equal to four days or more, intense menstrual bleeding, having premenstrual disorders, dysmenorrhea, or menstrual disorder leading to incapacity during the period, and using or not some medical method to reduce menstrual bleeding. About menstrual hygiene methods, in the last 12 months used external pads, internal or tampon pads, or menstrual cups; previous knowledge about all types of hygiene methods; whether knowledge was acquired by medical instruction or not, whether had previous use of the menstrual cup and if positive, the satisfaction level. On a scale from 1 to 10, we collected data about the influence of price, efficacy, sustainability, practicality, intimal health, menstrual hygiene, and preoccupation with internal genitalia manipulation on the choice of their preferred method.

The primary outcome variable was the menstrual cup’s actual use and preference as a menstrual hygiene method.

We used the statistical software package IBM SPSS Statistic version 24.0™ from IBM, Armonk, NY. The variable age was described as mean, standard deviation (SD), and Minimum / Maximum values and analyzed by independent sample Student’s t-test. A frequency described the categorical and binary variables. The dependent variable “use of menstrual cups,” which is categorical and binary, required a specific analysis, considering that it did not have a Normal distribution (which is an assumption for most analyzes). In this context, categorical data analysis allows us to use logistic regression to adapt traditional regression for categorical data. The clinical variables determined with the simple logistic regression analysis using a *P-value* <.200 were included in a multivariate logistic regression analysis. All significant variables identified by the simple logistic regression were used as predictors in the multivariate model. The relative risk (RR) and 95% confidence intervals (95%CI) were calculated for each variable. They were excluded one by one in the backward multivariate logistic regression analysis until reaching the best model, defined by the impossibility to exclude any other variable without significant loss in adjustment. For all multivariate logistic tests, the statistical significance limit was *P* <.05. We did not have missing data or incomplete questionnaires. We called the answer to the question “Do you use menstrual cups?” as a Dependent Variable and the other variables of interest as Predictor Variables since we tested the hypothesis that menstrual cups use depends on the other variables. The logistic regression model was conducted to identify the independent risk factors for the use of menstrual cups.

## Results

Of 277 included women, 164 participated, answering the online survey sent via text message cell phone in January 2021. The mean age was 22.26 years (SD: 3.21), with a minimum age of 18 and a maximum, 41 years. Table1 represents the background findings of the study. Table 2 shows the medical students’ menstrual cycle characteristics, and 82.32% had a regular cycle, lasting more than three days (78.05%), and 21.95% intense bleeding. Also, 98.78% had menstrual disorders and dysmenorrhea, and these factors are reported as disabling for 35.98% of women during the menstrual period. They reported the use of some method in order to reduce menstrual flow in 67.68% of participants. The menstrual hygiene products mentioned as used in the last 12 months were in order of frequency, the external absorbent or pad, the internal tampon, and the menstrual cup. Regarding knowledge about menstrual hygiene methods, 80.49% said they knew all the methods, and 25.60% said they had already received medical guidance about it. The concern with environmental biodegradation was reported by 68.29%. They have used the menstrual collector at some time, 22.56%. Of these, 17.07% reported some problem or difficulty with the method, and 5.49% showed dissatisfaction with the menstrual collector. Table 3 presents the items’ efficacy, hygiene, practicality, intimal health, price, sustainability, and integrity of the internal genitalia, when asked about on a scale from 1 to 10, where ten as a maximum score, as a denotation of powerful influence, and one as its minimum. Tables 4 and 5 show the results of the statistical analyses performed. Based on the results, the *P*-values < .20 were tested in the Multivariate regression model with the backward stepwise technique. Thus, the best model results in the variables with a *P*-value <.05 below. It was concluded that the factors of using any method to reduce menstruation in the last 12 months had an Adjusted Odds Ratio of 0.310 (95%CI 0.122-0.787), *P*-Value: .014. Concern about the environmental biodegradation caused by menstrual hygiene products had an Odds Ratio Adjusted of 6.369 (95%CI 1.372-29.562), *P*-value: .018; Intimal Health had an Odds Ratio Adjusted of 1.996 (95%CI 1.183-3.368), *P*-value: .010; internal genital integrity had an Odds Ratio Adjusted of 0.824 (95%CI 0.682-0.995), *P*-value: .045, for menstrual cups use.

**Table 1.** Sociodemographic characteristics (N=164)

|  |  |
| --- | --- |
| Age * | 22.26 (3.21)** |
| Caucasian | 148 (90.24%) |
| Not married | 158 (96.34%) |
| Catholic | 109 (66.46%) |
| Protestant | 22 (13.41%) |
| Another religion | 11 (6.71%) |
| No religion | 142 (86.59%) |

\*Age in Years.
\*\*Mean and Standard deviation
For the other variables, we used frequency and percentile.

**Table 2:** Menstrual and clinical variables. N=164.

| Variable | N | Percentile |
| --- | --- | --- |
| Irregular menstrual cycle | 29 | 17.68 |
| Menstrual period more than three days | 128 | 78.05 |
| Intense bleeding | 36 | 21.95 |
| Regular menses in the last 12 months | 110 | 67.07 |
| Dysmenorrhea or Menstrual Disorders | 162 | 98.78 |
| Incapacity due to menstrual period | 59 | 35.98 |
| Use of some method for reducing menstrual bleeding | 111 | 67.68 |
| Use of external pads in the last 12 months | 136 | 82.93 |
| Use of internal tampon in the last 12 months | 60 | 36.59 |
| Use of menstrual cups in the last 12 months | 28 | 17.07 |
| Amenorrhea in the last 12 months | 11 | 6.71 |
| Knowledge about all menstrual hygiene methods | 132 | 80.49 |
| Concern about biodegradation of menstrual hygiene methods | 112 | 68.29 |
| Medical orientation about menstrual cups | 42 | 25.60 |
| Previous use of menstrual cups | 37 | 22.56 |
| Any problem using menstrual cups | 28 | 17.07 |
| Insatisfaction with menstrual cups | 9 | 5.49 |

**Table 3:** Ordinal data concerning the influence of variables in choosing the last 12 months’ preferred hygiene method. N=164.

|  | Mean (SD) | Median * | Minimum- Maximum |
| --- | --- | --- | --- |
| Price | 6.27 (2.92) | 7 (5 - 8) | 1 - 10 |
| Efficacy | 9.56 (1.24) | 10 (10 - 10) | 1 - 10 |
| Sustainability | 6.15 (2.98) | 7 (4 - 8.25) | 1 - 10 |
| Practicality | 8.89 (1.84) | 10 (8 - 10) | 1 - 10 |
| Intimal Health | 8.80 (1.87) | 10 (8 - 10) | 1 - 10 |
| Hygiene | 9.42 (1.45) | 10 (10 - 10) | 1 - 10 |
| Internal genital Integrity | 2.76 (3.19) | 1 (1 - 3.25) | 1 - 10 |

\*(1st – 3rd Quartile)

**Table 4:** Simple regression model (*P*<.20) for each predictor variable. N= 164.

|  | Odds ratio | 95% Confidence Interval | *P*-value |
| --- | --- | --- | --- |
| Irregular periods | 0.311 | (0.069; 1.390) | 0.126 |
| More than three days of the menstrual period | 2.670 | (0.757; 9.413) | 0.127 |
| Intense menstrual bleeding | 0.964 | (0.358; 2.592) | 0.942 |
| A regular menstrual period in the last 12 months | 1.584 | (0.628; 3.997) | 0.330 |
| Incapacity during the menstrual period | 0.986 | (0.422; 2.305) | 0.975 |
| Any method to reduce menstrual bleeding | 0.402 | (0.176; 0.921) | 0.031 |
| Medical instructions about menstrual cups | 0.430 | (0.140; 1.321) | 0.141 |
| Biodegradation concerns | 7.558 | (1.721; 33.198) | 0.007 |
| Price | 1.111 | (0.956; 1.290) | 0.168 |
| Efficacy | 1.179 | (0.746; 1.863) | 0.481 |
| Sustainability | 1.413 | (1.172; 1.705) | < 0.001 |
| Practicability | 1.582 | (1.023; 2.445) | 0.039 |
| Hygiene | 1.977 | (1.174; 3.331) | 0.010 |
| Intimal Health | 1.496 | (0.846; 2.645) | 0.166 |
| Internal genital integrity | 0.850 | (0.709; 1.020) | 0.166 |

\* Dysmenorrhea or Menstrual Disorders: No sufficient sample representation
\*\*Using no as reference.
$P<.20$

**Table 5:** The predicted variables of the best model of multiple logistic regression. N=164.

|  | Odds ratio<br>adjusted | 95%<br>Confidence<br>Interval | *P*-<br>value |
| --- | --- | --- | --- |
| Any method to reduce menstrual<br>bleeding | 0.310 | (0.122; 0.787) | .014 |
| Biodegradation concerns | 6.369 | 1.372; 29.562) | .018 |
| Intimal Health | 1.996 | (1.183; 3.368) | .010 |
| Internal genital integrity | 0.824 | (0.682; 0.995) | .045 |
| *Backward stepwise method*<br>*P*<.05 |  |  |  |

## Discussion

The menstrual cup method was previously used by 22.56% of women students from a private medical school in Brazil. We found significant evidence that women have not used some method to reduce menstrual flow, women concerned about the biodegradation of personal hygiene products, women concerned about their intimal health, and women not bothered by the manipulation of their internal genitalia are the characteristics that influenced positively in the choice of the menstrual cup method.

Women concerned about environmental degradation have a 536.9% greater chance of choosing menstrual cups. This may be related to the lesser amount of plastic waste resulting from a menstrual cup device in 10 years compared to disposable methods, being that the menstrual cups represent 6% of plastic waste compared to the residues of the internal absorbents and 0.4% of the external absorbents. Intimal health was the second most crucial factor in choosing the product in our study. Although the use of the menstrual cup requires manipulation of the genitalia, evidence shows that after the correct instruction and an adaptation time of an average of 6 months, there is a good acceptance process. (van Eijk et al. 2018) Therefore, the negative effect on the choice of the menstrual cup by women who are concerned with manipulating the genitalia, evidenced in our study and another study carried out in Zimbabwe, can be conducted by the proper instruction concerning the anatomy of the internal genitalia, and on the correct form of insertion, without impairing physical integrity, even in virgin women. (Tembo et al. 2020)

The knowledge and access to most menstrual hygiene products, reported by more than 80.49% of the participants, were not an expected factor influencing the choice of the menstrual cups in this study. Otherwise, 19.51% are still unaware of some products available, which is similar to that found in a study by van Erjk in high-income countries. (van Eijk et al. 2019)

## Limitations

the primary limitation is that the exposure and outcome were simultaneously assessed in the study’s cross-sectional design. We had good medical students’ compliance and a high degree of understanding of this population’s questions compensated for a non-validated questionnaire. Moreover, there is a lack of validated questionnaires in the literature about the proposed theme. Nevertheless, we believe that the results obtained are relevant and can be generalized to the women’s population, especially those with higher educational levels.

## Conclusion

The menstrual cup is a safe and practice method for menstrual hygiene, underused by women, even in a population with evident knowledge and access to menstrual hygiene options. We observed that women using something to reduce their menstrual flow, women concerning biodegradation of menstrual hygiene products, women concerning intimal health, and those with no concerns about manipulating their internal genitalia were independent factors for the adherence of the menstrual cup. Further studies are necessary to investigate the mechanisms involved in hygiene menstrual methods’ preferences and choices and promote interventions to enforce menstrual cups among women.

## Data Availability

All data produced in the present study are available upon reasonable request to the authors

## Acknowledgments

Special consideration for the support from FEMA Medical Faculty and Mr. Robin O’Brien, Managed Services Consultant, for the English review.

## References

Eijk, Anna Maria van, Kayla F. Laserson, Elizabeth Nyothach, Kelvin Oruko, Jackton Omoto, Linda Mason, Kelly Alexander, et al. 2018. “Use of Menstrual Cups among School Girls: Longitudinal Observations Nested in a Randomised Controlled Feasibility Study in Rural Western Kenya.” Reproductive Health 15 (1): 139. https://doi.org/10.1186/s12978-018-0582-8.

Eijk, Anna Maria van, Garazi Zulaika, Madeline Lenchner, Linda Mason, Muthusamy Sivakami, Elizabeth Nyothach, Holger Unger, Kayla Laserson, and Penelope A. Phillips-Howard. 2019. “Menstrual Cup Use, Leakage, Acceptability, Safety, and Availability: A Systematic Review and Meta-Analysis.” The Lancet Public Health 4 (8): e376–93. https://doi.org/10.1016/S2468-2667(19)30111-2.

Elm, Erik von, Douglas G. Altman, Matthias Egger, Stuart J. Pocock, Peter C. Gøtzsche, Jan P. Vandenbroucke, and STROBE Initiative. 2008. “The Strengthening the Reporting of Observational Studies in Epidemiology (STROBE) Statement: Guidelines for Reporting Observational Studies.” Journal of Clinical Epidemiology 61 (4): 344–49. https://doi.org/10.1016/j.jclinepi.2007.11.008.

Hennegan, Julie, Agnes Nansubuga, Calum Smith, Maggie Redshaw, Agnes Akullo, and Kellogg J. Schwab. 2020. “Measuring Menstrual Hygiene Experience: Development and Validation of the Menstrual Practice Needs Scale (MPNS-36) in Soroti, Uganda.” BMJ Open 10 (2): e034461. https://doi.org/10.1136/bmjopen-2019-034461.

Jahan, Farjana, Md Nuruzzaman, Farhana Sultana, Mehjabin Tishan Mahfuz, Mahbubur Rahman, Farhana Akhand, Stephen P. Luby, Leanne Unicomb, and Peter J. Winch. 2020. “Piloting an Acceptable and Feasible Menstrual Hygiene Products Disposal System in Urban and Rural Schools in Bangladesh.” BMC Public Health 20 (1). https://doi.org/10.1186/s12889-020-09413-x.

North, Barbara B., and Michael J. Oldham. 2011. “Preclinical, Clinical, and Over-the-Counter Postmarketing Experience with a New Vaginal Cup: Menstrual Collection.” Journal of Women’s Health 20 (2): 303–11. https://doi.org/10.1089/jwh.2009.1929.

Smith, Annie D., Alfred Muli, Kellogg J. Schwab, and Julie Hennegan. 2020. “National Monitoring for Menstrual Health and Hygiene: Is the Type of Menstrual Material Used Indicative of Needs across 10 Countries?” International Journal of Environmental Research and Public Health 17 (8): 2633. https://doi.org/10.3390/ijerph17082633.

Tembo, Mandikudza, Jenny Renju, Helen A. Weiss, Ethel Dauya, Tsitsi Bandason, Chido Dziva-Chikwari, Nicol Redzo, et al. 2020. “Menstrual Product Choice and Uptake among Young Women in Zimbabwe: A Pilot Study.” Pilot and Feasibility Studies 6 (1): 182. https://doi.org/10.1186/s40814-020-00728-5.

